# Predicting Behavioral Determinants of Health from Clinical Text Using Transformer Models and BiLSTM

**DOI:** 10.1101/2025.10.13.25337944

**Authors:** Saad Althabiti, Chuming Chen, Cathy Wu, Vijay Shanker

## Abstract

**Objective:** Social and behavioral determinants of health play a critical role in patient outcomes, yet much of this information is documented only in unstructured clinical text rather than structured records. Detecting these factors using natural language processing enables a deeper understanding of patient health and supports better decision-making. This study aims to improve the prediction of Behavioral Determinants of Health (BDoH) from medical records by systematically comparing multiple transformer-based models: Bio-ClinicalBERT, BioBERT, BioMedBERT, RoBERTa, and T5-EHR with and without BiLSTM. We also address the challenge of class imbalance in the dataset and assess the role of generative versus discriminative models in this task.

**Methods:** We evaluated five transformer models in combination with BiLSTM for classifying BDoH mentions in the MIMIC-III dataset. To address the challenge of class imbalance, we compared oversampling, undersampling, and class weighting strategies to identify the most effective approach. Then, each model was first tested as a standalone classifier to establish baseline performance. We then extracted embeddings from these models and used them as input to a BiLSTM layer to investigate whether sequential modeling could further improve classification. Also, by introducing our T5-EHR model alongside BERT and RoBERTa models, we were able to assess both the benefits of BiLSTM and the value of using a generative model for this task. Finally, we benchmarked the performance of the best approaches against the published results from the MIMIC-SBDH study, using precision, recall, and F1 scores as evaluation metrics.

**Results:** The experiments showed that class weighting was the most effective strategy for handling class imbalance, showing better performance across all models. The addition of BiLSTM improved the performance of Bio-ClinicalBERT, BioMedBERT, and BioBERT across all categories. RoBERTa and T5-EHR achieved strong results as standalone models, with no additional benefit from BiLSTM integration. Across all evaluated labels, T5-EHR achieved the highest F1 scores compared to the other models and prior baselines in this study.

**Conclusion:** This study demonstrates that handling class imbalance is essential for robust prediction of social and behavioral determinants of health from clinical notes. We find that the integration of BiLSTM provides clear benefits for Bio-ClinicalBERT and BioBERT, enhancing their ability to capture sequential dependencies. However, RoBERTa and T5-EHR achieved their best performance without BiLSTM, reflecting the strength of their pretrained representations. Among all models, our pretrained T5-EHR, adapted especially for clinical text, outperformed every baseline, establishing the highest F1 scores across all labels and surpassing the results reported in the MIMIC-SBDH benchmark. These findings highlight both the value of model-specific strategies and the effectiveness of generative models for advancing state-of-the-art classification in health informatics.

## Introduction

The World Health Organization defines social determinants of health (SDOH) as the conditions in which people are born, grow, live, work, and age, influenced by factors such as education, employment, and social support, shaped by money, power, and resources [1]. Behavioral determinants include drug, alcohol, and tobacco use, which directly impact health and can be modified through interventions. Social and behavioral determinants of health (SBDHs) are crucial in shaping health outcomes, influencing behaviors, treatment adherence, and well-being. Understanding SBDHs is essential for developing effective health policies and interventions to improve population health and reduce disparities.

SBDHs factors are often found in the social history section of clinical notes, which includes key information about social, behavioral, and environmental determinants of health [2]. This section contains rich qualitative data that provides insights into the patient’s life circumstances and behaviors not captured in structured data fields. Accurate extraction and analysis of these factors are essential for comprehensive patient care and health outcome prediction. Understanding these determinants allows healthcare providers to create more personalized and effective care plans. Therefore, NLP techniques that extract SBDHs factors from unstructured clinical text are crucial, enabling systematic analysis of large volumes of text to provide insights that inform clinical decision-making and public health strategies.

Although transformer-based models BERT and BiLSTM hybrids method have shown success in NLP, a comprehensive evaluation of their combined and standalone use in the clinical domain, particularly for extracting SBDHs from unstructured text, is still lacking, motivating our work. Our work addresses this gap by systematically comparing a set of widely used domain-specific BERT models (Bio-ClinicalBERT [3], BioBERT [4], BioMedBERT [5]), RoBERTa [6] as an improved version of BERT, and our pretrained generative model (T5-EHR), with and without BiLSTM [7]. We focus specifically on the behavioral aspects of SBDHs because these factors play a critical role in patient outcomes but are not reported in structured data and only documented in free-text clinical notes. Behaviors such as smoking, alcohol consumption, and substance use are strongly linked to chronic disease, accounting for a substantial proportion of preventable morbidity and mortality worldwide. This study aims to advance NLP methods for extracting SBDH factors from the MIMIC-III dataset by addressing the challenge of class imbalance, investigating the role of BiLSTM in enhancing sequential modeling, and exploring the potential of domain-adapted generative models. Our goal is to enable more comprehensive patient risk assessment and support population health research.

Our study contributes not only by providing a systematic evaluation of transformer-based models for extracting SBDHs from unstructured clinical text, but also by investigating techniques to improve current state-of-the-art results. We extend the scope of analysis by addressing the challenge of class imbalance, examining the added value of sequential modeling through BiLSTM, and exploring the potential of generative models with our T5-EHR model, while directly benchmarking against the study referenced in [8]. These contributions are guided by the following research questions:

1. Which class imbalance strategy provides the most effective performance across transformer-based models for SBDH classification?
2. What is the contribution of incorporating BiLSTM in BERT-based modeling for improving sequential modeling of clinical text?
3. How does our pretrained generative model, T5-EHR, compare to the best-performing approaches from prior experiments?
4. To what extent does our proposed approaches improve upon the state-of-the-art results reported in the MIMIC-SBDH benchmark [8]?

### Related Work

There are limited studies investigating NLP methods, including the use of BERT [9] and BiLSTM [10], to extract SBDHs information from clinical text data. The main challenges include the lack of annotated training data, the complexity and variability of the text in health records, and the need for substantial manual effort to annotate such data. Also, patients’ privacy and data confidentiality limit access to medical records. Despite these challenges, some efforts have been made to address this gap. For example, [11] aimed to detect the presence of SBDHs in patient records and identify 11 SBDH categories using data from Columbia University Medical Center. They tested classifiers like Logistic Regression, SVM, Random Forests, CaRT, and AdaBoost. This study demonstrates the potential of machine learning for extracting SBDHs and the need for advanced NLP methods suited to clinical text complexities.

Another study [12] used five classifiers, Linear SVM, K-Nearest Neighbors, Random Forest, XGBoost, and BiLSTM, to determine if six SBDH topics were documented in clinical notes from the UNC clinical data warehouse. This highlights the applicability of both traditional and deep learning methods in SBDH extraction. Additionally, [8] developed a publicly available dataset with annotations using MIMIC III, focusing on the status of seven SBDHs and employing Random Forest, XGBoost, and Bio-ClinicalBERT. The annotated categories include four social factors (Community, Education, Economics, and Environment) and three behavioral factors (Tobacco Use, Alcohol Use, and Drug Use). Their use of Bio-ClinicalBERT highlights the value of domain-specific language models for enhancing SBDH extraction from clinical notes. In the present work, we use this same MIMIC-SBDH dataset as the foundation for all our experiments and model evaluations. Additionally, [13] used the open-source NLP system NimbleMiner to extract alcohol and substance abuse status from the MIMIC III dataset, finding that the Random Forest algorithm achieved the highest F-score. Another study [14] focused on SBDH concepts in cancer patient records, annotating five concepts in EHR data from the UF Health Integrated Data Repository. They evaluated BERT, RoBERTa [15], and LSTM [10], showing that transformer-based models outperformed LSTM in extracting meaningful SBDH information. Other tools, such as MTERMS [16], have also been used to extract social factors from physician notes [17].

Another effort [18] aimed to develop a framework for automated classification of multiple SDOH categories using the MIMIC-III dataset. They curated thirteen SDOH categories and established annotation guidelines to ensure consistency and accuracy, resulting in 3,504 annotated sentences from the “Social Work” category in MIMIC-III clinical notes. However, this annotated dataset is not publicly available. They also tested three deep neural network architectures, CNN, LSTM, and BERT, for automated detection of eight SDOH categories.

Moreover, some studies have focused on specific patient groups to address unique challenges associated with their conditions. For example, [19] developed a rule-based NLP algorithm to identify seven SDOH domains, such as housing, transportation, food and medication insecurities, social isolation, abuse, neglect, exploitation, and financial difficulties, for patients with Alzheimer’s disease and related dementias (ADRD) using unstructured EHR data from the Michigan Medicine database. They compared the rule-based method with deep learning and regularized logistic regression approaches. This targeted approach helps in understanding and addressing the social challenges faced by ADRD patients, thereby enhancing their care and quality of life.

Another study [20] addressed three key limitations in SDOH extraction: identifying multiple SDOH events of the same type within a sentence, handling overlapping SDOH attributes, and capturing SDOH spanning multiple sentences. These challenges affect the comprehensive extraction of SDOH information, limiting NLP models’ effectiveness. Their work involved two stages: first, fine-tuning a Bio-ClinicalBERT-based NER system to extract SDOH event triggers, and second, using a multitask, multilabel NER to extract arguments for these events. This two-stage method enhances the precision and contextual understanding of SDOH events. The data used was from the 2022 n2c2 Track 2 competition, which includes clinical notes annotated for SDOH. In this study, [21] introduced two main contributions: mSpERT, an advanced deep learning model for SDOH extraction, trained on the Social History Annotated Corpus (SHAC) from the 2022 National NLP Clinical Challenges SDOH task, and a large-scale EHR case study. mSpERT accurately identifies SDOH events from clinical narratives, improving extraction precision. The case study applied mSpERT to 225,089 patients and 430,406 clinical notes from the University of Washington Medicine, showing that integrating SDOH data from narratives with structured EHR data provides a fuller picture of patient health. This approach aids in better care, risk assessment, and identifying social needs by filling gaps left by structured data alone. Additionally, this study [22] introduces a method combining BERT embeddings with BiLSTM to improve emotion analysis in dialogue systems, addressing challenges like short text and reversed word order. By using BERT for word-level and sentence-level vectors and BiLSTM for bidirectional semantic dependencies, the model enhances emotion classification accuracy.

Finally, two more work investigate the effectiveness of combining BERT and BiLSTM, but for the task of sentiment analysis. This study [23] proposes a hybrid model for sentiment classification based on BERT, BiLSTM, and TextCNN to address challenges in analyzing netizens’ comments, such as short text, open vocabulary, and reversed word order. The model leverages BERT for contextual embeddings, BiLSTM for capturing sequential dependencies, and TextCNN for extracting local features. Similarly, this study [24] proposes using BERT combined with CNN, RNN, and BiLSTM for sentiment analysis of tweets, aiming to address challenges like varying text lengths and ambiguous emotional information.

## Methods

In this section, we will first describe the dataset used in our investigation and highlight its characteristics. We will also describe the class imbalance issue in the dataset. Finally, we will describe the models we consider as well as the evaluation metrics that are reported in this project.

### Dataset

We obtained the annotation information from MIMIC-SBDH [8], which is one of the publicly available resources for studying SBDH extraction. In this project, they used discharge summaries from MIMIC-III (Medical Information Mart for Intensive Care III) dataset [25]. MIMIC-III is a large, freely available database comprising de-identified health-related data associated with over forty thousand patients who stayed in critical care units of the Beth Israel Deaconess Medical Center between 2001 and 2012. The dataset has the SBDHs information under social history section in the discharge summaries. Additionally, the database includes a wide range of data types, such as demographics, vital sign measurements, and laboratory test results. This extensive variety of data provides a comprehensive view of patient health, making it an invaluable resource for research on social and behavioral determinants of health.

In this dataset, 7,025 discharge summaries containing social history sections were randomly selected for annotation, ensuring a diverse and representative sample. Moreover, each behavioral factor was labeled with one of five labels: Present, Past, Never, Unsure, and None, as defined in Table 1^1^. These labeling rules ensure consistent and clear categorization of behavioral factors across all records. More details about the annotation can be found in their published work. This detailed annotation process is crucial for developing accurate NLP models that can reliably extract and classify SBDH information from unstructured clinical text, providing a solid foundation for further research and model development. Figure 1 shows the class distribution for Behavioral Determinants of Health.

**Table 1:** Labels definitions (Tobacco use)

| Label | Definitions |
| --- | --- |
| Present | If the patient was a current consumer of tobacco. |
| Past | If the patient was a consumer in the past and had quit. |
| Never | If the patient had never consumed tobacco. |
| Unsure | If the discharge summary had an ambiguous passage related to tobacco consumption |
| None | If there was no related text. |

**Figure 1:**
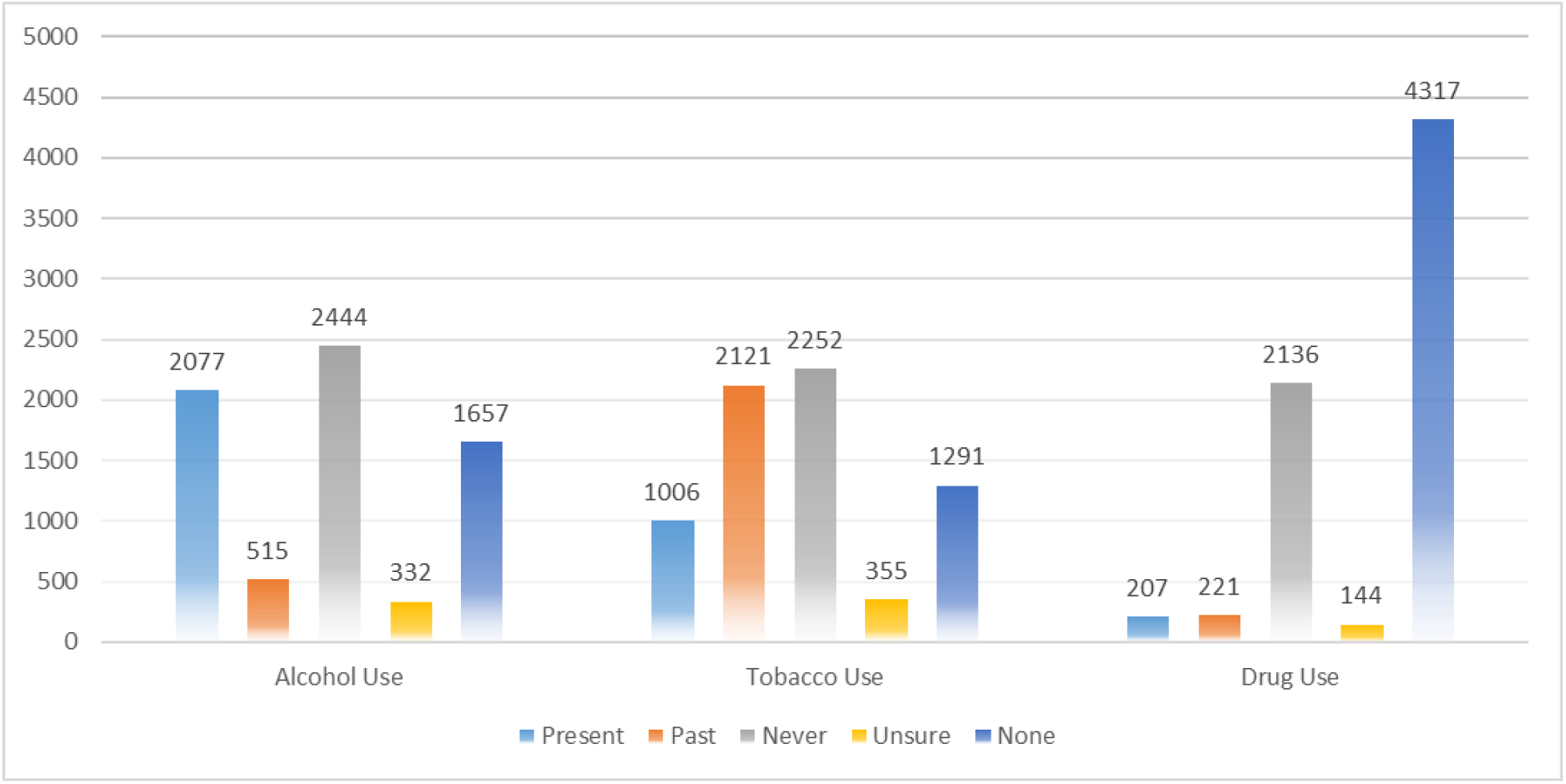
Class Distribution for Behavioral Determinants of Health

The MIMIC-SBDH dataset includes three behavioral determinants of health (Tobacco Use, Alcohol Use, and Drug Use), and four social determinants of health (Community, Education, Economics, and Environment). In this work, we focus primarily on the three behavioral categories because these factors are strongly and directly associated with adverse health outcomes, such as cardiovascular disease, cancer, and mental health disorders, and they play a critical role in disease prevention and patient management. Understanding and predicting behavioral determinants can therefore have immediate clinical relevance for intervention planning. However, for completeness, we also evaluate our models on the four social categories and compare their performance with the results reported in the original study.

Class imbalance is a significant challenge in this dataset, particularly for the Drug Use label. As shown in Figure 1, The Drug Use label is highly imbalanced because mentions of drug use are relatively rare in clinical notes. This imbalance can cause models to favor the majority class, leading to missed detections of other labels. Clinically, identifying patients with drug use history is important due to its strong link to adverse outcomes and treatment complications. Addressing this imbalance is therefore essential for improving both model fairness and clinical service.

### Models

In this project, we designed a series of experiments to evaluate different modeling strategies for classifying SBDHs. We first compared several transformer models as standalone classifiers. These models differ in the model pretraining domain, whether they include a specialized vocabulary, and training objective. We then extended these experiments by adding a BiLSTM layer for sequential processing during fine-tuning. Finally, we evaluated our pretrained generative model, T5-EHR, both with and without BiLSTM, enabling a comprehensive comparison across discriminative and generative architectures. We evaluated these models in this study:

#### Bio-Clinical BERT [3]

This model was initialized from BioBERT and further trained using clinical texts from the approximately 2 million notes in the MIMIC-III dataset. This model is considered the first public model that was released for the clinical domain and clinical NLP tasks [3]. Bio-ClinicalBERT leverages its extensive pretraining on clinical notes to excel in tasks requiring deep understanding of clinical language and patient history, making it particularly suitable for our experiments.

#### BioBERT [4]

It is the first domain-specific BERT based model that pretrained on large-scale biomedical corpora. This model achieved high scores in biomedical downstream tasks such as named entity recognition, relation extraction, and question answering tasks. The full model was pretrained on English Wikipedia, books corpus, PubMed abstracts, and PMC full text articles [4]. BioBERT’s pretraining on biomedical-specific texts provides it with a superior understanding of medical terminology and context, which is essential for accurately predicting SBDH factors.

#### BioMedBERT (Previously known as PubMedBERT) [5]

Unlike previous biomedical BERT models that rely on continual pretraining from general-domain BERT, BioMedBERT was trained from scratch using only PubMed abstracts, with a vocabulary derived entirely from biomedical text. This domain-specific pretraining strategy addresses the limitations of mixed-domain approaches, where general-domain vocabularies often fragment medical terms into irrelevant sub-words. BioMedBERT demonstrated consistent improvements across a wide range of biomedical NLP tasks, including NER, relation extraction, sentence similarity, document classification, and question answering.

#### RoBERTa-base-PM-M3-Voc [6]

RoBERTa is an improved version of BERT that optimizes pretraining by removing the next-sentence prediction objective, using dynamic masking, and training on much larger batches and corpora. The variant we used is pretrained on a combination of PubMed abstracts, PMC full-text articles, and MIMIC-III clinical notes, with domain-specific vocabulary (M3-Voc) designed for biomedical and clinical language. This adaptation enables the model to better capture domain terminology and context, allowing it to perform strongly on both biomedical and clinical NLP tasks.

#### T5-EHR

It is our domain-specific generative model, adapted from the Text-to-Text Transfer Transformer (T5) framework. We developed thirteen different pretrained variants of T5-EHR using different combinations of biomedical and clinical corpora, including MIMIC-III and MIMIC-IV notes, PubMed abstracts, and PMC articles. These models are currently under revision for public release due to the privacy restrictions of the MIMIC datasets. Unlike discriminative models such as BERT or RoBERTa, T5-EHR frames all tasks in a text-to-text format, enabling a flexible approach to classification. Based on our evaluation, the variant pretrained solely on MIMIC with a MIMIC-derived vocabulary achieved the highest F1 score on the Drug Use label, which is the most imbalanced and challenging category. Therefore, we selected this variant for the remaining experiments in this study.

#### Bidirectional Long Short-Term Memory Network (BiLSTM) [7]

BiLSTM is a type of neural network architecture designed to understand and analyze sequences of data, such as sentences in natural language. BiLSTM processes the input sequence in both forward and backward directions simultaneously. This approach captures the long-range dependencies and context.

As part of our experiments, we incorporated a BiLSTM layer on top of the transformer embeddings to evaluate its impact on sequence modeling. The integration of BiLSTM allows the model to capture sequential dependencies and contextual information more effectively, leading to better performance by leveraging the sequential relationships within the text. Figure 2 illustrates this architecture. The final hidden state from the BiLSTM layer is passed to the classification layer, which captures the overall context of the text. A dense layer with a softmax activation then converts this representation into probabilities for each label.

**Figure 2:**
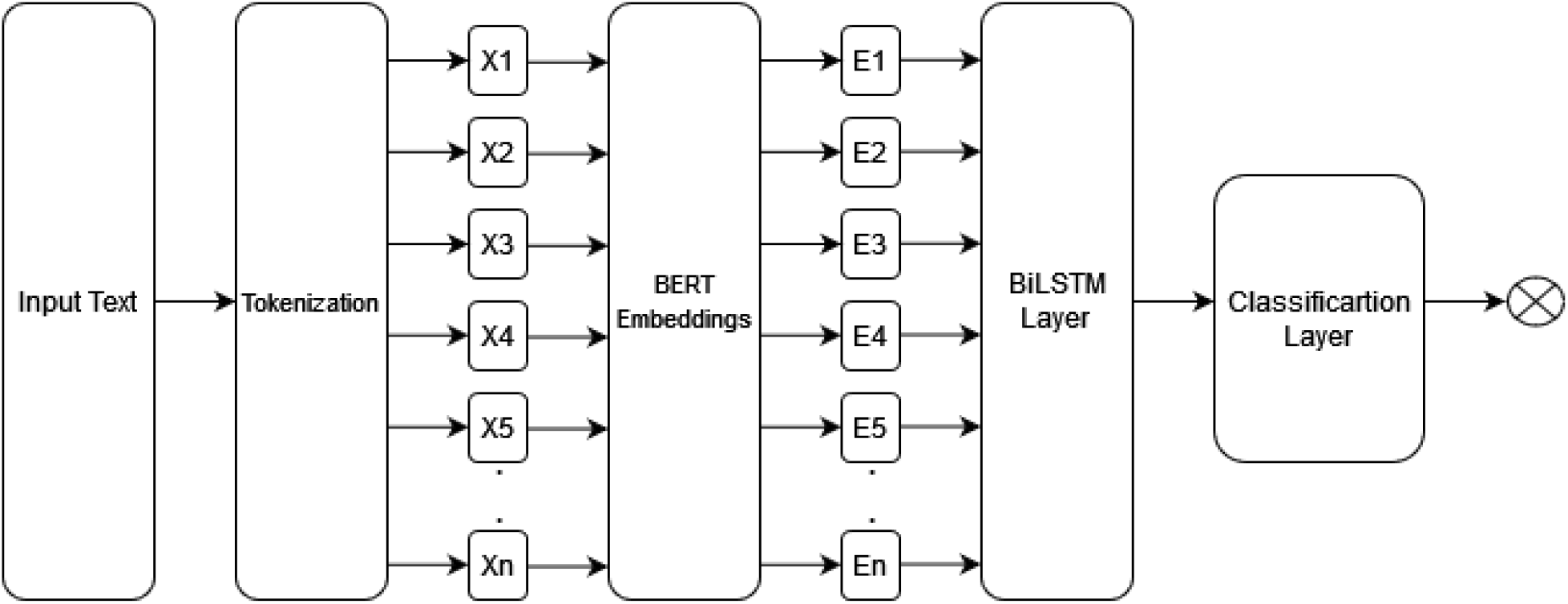
BiLSTM-BERT Architecture

For comparison, we also implemented a baseline model using the transformer alone, where the [CLS] token embedding was connected directly to a dense layer and softmax classifier for prediction. Figure 3, obtained from [26], illustrates this architecture.

**Figure 3:**
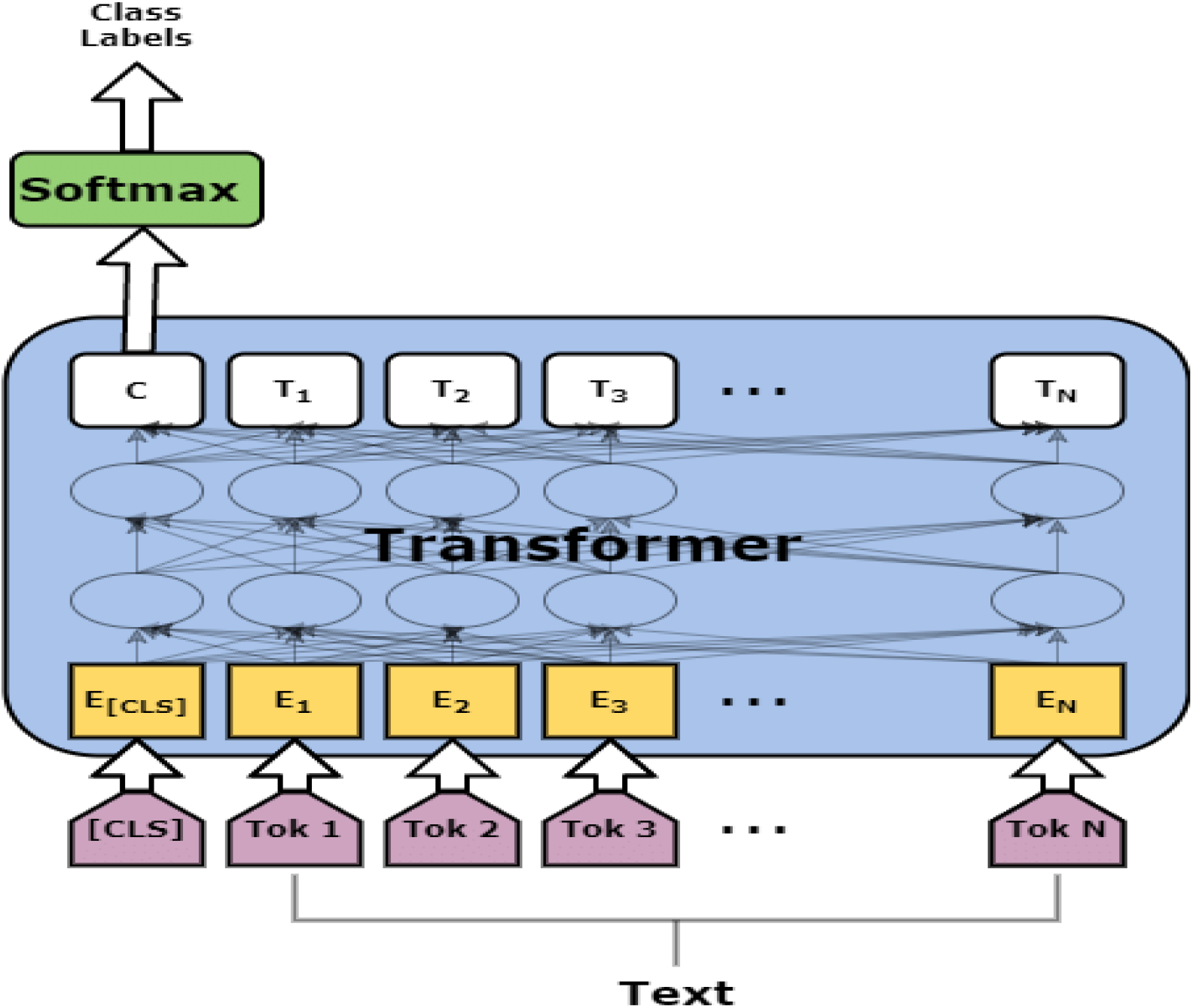
Architecture of BERT Models for Text Classification, Obtained From [26]. In this work, N = 510, represents the number of tokens between the special [CLS] and [SEP] tokens, with a maximum sequence length of 512.

### Class Imbalance

The dataset has a significant class imbalance in all labels as shown in Figure 1. This imbalance can lead to biased model predictions, favoring the majority classes and neglecting the minority ones, which is particularly problematic in clinical applications where accurate identification of all classes is crucial for patient care and treatment decisions. To resolve this problem, we calculated class weights using the *compute_class_weight* function from *scikit-learn*. This method assigns higher weights to underrepresented classes and lower weights to overrepresented ones, based on the distribution of labels in the training dataset. Specifically, the weights are calculated to ensure that each class has a balanced influence during model training. The formula considers the inverse of the class frequencies, so minority classes receive higher weights, helping to mitigate the effects of class imbalance and reducing bias towards the majority classes. We used those weights in the *CrossEntropyLoss* function that computes the cross-entropy loss between input logits and target. By incorporating class weights into the loss function, we ensure that the model learns to treat each class according to its importance, thus improving the overall balance of predictions.

Additionally, we evaluated oversampling and undersampling methods. Randomly oversampling involves duplicating instances of the minority classes to balance the class distribution, while randomly undersampling involves reducing the number of instances in the majority classes to achieve a similar balance. These methods modify the dataset to create a more balanced distribution of classes, which help improve the model’s ability to learn from minority class examples. In our experiments, the class weight method had the highest F1 score, and it was the best option for us since we do not have to remove or duplicate any records from the dataset. This method preserves the original dataset’s integrity while effectively addressing the class imbalance issue, leading to more reliable and generalized model performance.

### Experiment Setups and Evaluation Methods

We extracted the social history from each discharge summary (7,025 records) and used it as input to our models. All non-alphanumeric characters were removed from the texts. Also, the texts were lower-cased, and a maximum sequence length of 512 tokens was set. This preprocessing step helps to eliminate noise and irrelevant characters, ensuring that the input data is clean and standardized, which is essential for effective model training. After that, we tested the following common parameters in practice: Epochs: [20], Batch Sizes: [8,16, 32], Learning Rates: [0.1, 0.01, 1e-3, 1e-4, 5e-5, 5e-6], Number of Unites in Layer 1: [64, 128, 256] Number of Unites in Layer 2: [64, 128, 256]. These values were selected due to their common use in similar models and their ability to explore a broad range of training conditions.

Then, we split the provided dataset into training (80%), development (10%), and test (10%) sets using a random seed of 42. The development dataset was used to determine the optimal parameters for each experiment. After testing different learning rates, a learning rate of 5e–5 proved to be optimal for the BERT-based models and was used consistently across all of them. When integrating BiLSTM, a smaller learning rate of 5e–6 provided the best performance. For the generative model T5-EHR, the same learning rate produced suboptimal results, and after additional testing, a learning rate of 1e–4 was found to be optimal. Finally, we fine-tuned all the experiments for 20 epochs using Adam optimizer. A batch size of 32 was used for all experiments and the test set was then used for the final evaluation.

To evaluate our results, we reported macro-averaged precision, recall, and F1 score for each experiment. For this project, we utilized an A100 Nvidia GPU on the Google Colab+ platform to fine-tune the models for the classification tasks, leveraging the GPU’s computational power to handle the extensive training processes required for deep learning models. Using the A100 Nvidia GPU significantly reduced the training time and allowed for more complex models to be trained efficiently. The training duration using only the BERT models is less than two minutes per epoch. Therefore, in our experiments, the total training time for 20 epochs is 34.5 minutes per category (e.g., Drug Use). In contrast, when incorporating BiLSTM, the training time for 20 epochs increases to 50.8 minutes per category, equating to approximately 2.5 minutes per epoch.

## Results

### Effectiveness of Class Imbalance Strategies

At first, we evaluated different methods for handling the imbalance data, which is a critical issue that can skew the model’s performance, leading to biased predictions favoring the majority class. Table 2 shows a detailed assessment of these methods. In this experiment, “drug use” label was used to evaluate the methods because it represents the most imbalanced category in the dataset. This label provided a challenging test case for assessing the effectiveness of different imbalance management techniques. We used ten epochs in this assessment since the goal of this step was to compare imbalance handling strategies rather than to fully fine-tuning the models.

**Table 2:** Imbalance Data Methods Comparison (Drug Use)

| Methods | Bio-ClinicalBERT |  |  | BioBERT |  |  | BioMedBERT |  |  | RoBERTa |  |  | T5-EHR |  |  |
| --- | --- | --- | --- | --- | --- | --- | --- | --- | --- | --- | --- | --- | --- | --- | --- |
|  | *P* | *R* | *F1* | *P* | *R* | *F1* | *P* | *R* | *F1* | *P* | *R* | *F1* | *P* | *R* | *F1* |
| No Method | 0.49 | 0.50 | 0.49 | 0.53 | 0.53 | 0.52 | 0.60 | 0.53 | 0.50 | 0.72 | 0.74 | 0.73 | 0.77 | 0.74 | 0.75 |
| Over-sampling | 0.53 | 0.60 | 0.55 | 0.53 | 0.64 | 0.56 | 0.47 | 0.59 | 0.50 | 0.69 | 0.71 | 0.70 | 0.77 | 0.74 | 0.75 |
| Under-sampling | 0.50 | 0.65 | 0.54 | 0.49 | 0.68 | 0.53 | 0.56 | 0.52 | 0.52 | 0.55 | 0.74 | 0.60 | 0.37 | 0.42 | 0.37 |
| Class Weights | **0.55** | **0.66** | **0.58** | **0.61** | **0.68** | **0.62** | **0.53** | **0.63** | **0.56** | **0.76** | **0.74** | **0.75** | **0.79** | **0.74** | **0.76** |

Based on the results, we decided to choose the class weight method in the following experiments. This approach provides best results and avoids any data manipulation. This approach also allows the model to pay more attention to underrepresented classes by adjusting the loss function, thereby enhancing the model’s ability to learn from minority class examples without altering the actual data distribution.

### Contribution of BiLSTM to Transformer Embeddings

Next, we examined the contribution of BiLSTM when added on top of transformer embeddings. This experiment was designed to measure whether BiLSTM provides additional benefits in capturing sequential dependencies. To do this, we first evaluated all models in their standalone form, where each transformer is paired with a simple classification head. These results are summarized in Table 3, which establishes the baseline performance of Bio-ClinicalBERT, BioBERT, BioMedBERT, and RoBERTa-base-PM-M3-Voc, all fine-tuned using the class-weighting strategy to address class imbalance.

**Table 3:** Standalone Transformer Models with Class Weighting for SBDH Classification.

| SBDH | Bio-ClinicalBERT |  |  | BioBERT |  |  | BioMedBERT |  |  | RoBERTa |  |  |
| --- | --- | --- | --- | --- | --- | --- | --- | --- | --- | --- | --- | --- |
|  | *P* | *R* | *F1* | *P* | *R* | *F1* | *P* | *R* | *F1* | *P* | *R* | *F1* |
| Alcohol Use | 0.75 | 0.78 | 0.76 | 0.73 | 0.77 | 0.74 | 0.67 | 0.73 | 0.69 | 0.89 | 0.87 | 0.87 |
| Tobacco Use | 0.80 | 0.77 | 0.77 | 0.85 | 0.81 | 0.83 | 0.75 | 0.79 | 0.76 | 0.90 | 0.90 | 0.90 |
| Drug Use | 0.54 | 0.63 | 0.57 | 0.57 | 0.67 | 0.60 | 0.57 | 0.66 | 0.58 | 0.76 | 0.74 | 0.75 |

As shown in Table 3, RoBERTa achieved the best overall results across all categories. BioBERT performed well, followed by Bio-ClinicalBERT and BioMedBERT. The Drug Use label remained the most challenging due to severe class imbalance. We then extended the evaluation by feeding the embeddings from each model into a BiLSTM layer to test whether sequential modeling provides measurable improvements. The results of this setup are presented in Table 4

**Table 4:** Transformer Embeddings Combined with BiLSTM for SBDH Classification.

| **SBDH** | **Bio-ClinicalBERT + BiLSTM** |  |  | **BioBERT + BiLSTM** |  |  | **BioMedBERT + BiLSTM** |  |  | **RoBERTa + BiLSTM** |  |  |
| --- | --- | --- | --- | --- | --- | --- | --- | --- | --- | --- | --- | --- |
|  | *P* | *R* | *F1* | *P* | *R* | *F1* | *P* | *R* | *F1* | *P* | *R* | *F1* |
| Alcohol Use | 0.83 | 0.86 | 0.84 | 0.80 | 0.85 | 0.82 | 0.77 | 0.76 | 0.76 | 0.57 | 0.43 | 0.34 |
| Tobacco Use | 0.88 | 0.89 | 0.88 | 0.90 | 0.87 | 0.88 | 0.85 | 0.82 | 0.83 | 0.66 | 0.56 | 0.56 |
| Drug Use | 0.75 | 0.74 | 0.74 | 0.74 | 0.72 | 0.72 | 0.72 | 0.67 | 0.69 | 0.38 | 0.38 | 0.37 |

As shown, Bio-ClinicalBERT, BioBERT, and BioMedBERT benefited from BiLSTM integration, achieving higher F1 scores across labels. In contrast, RoBERTa produced the weakest results overall, and its performance did not improve with BiLSTM integration, indicating that additional sequential modeling did not benefit the model for this task compared to domain-specific BERT models. This two-step comparison highlights that the usefulness of BiLSTM depends on the selected model for the targeted task.

### Comparison of T5-EHR to Prior Best-Performing Approaches

We then compared our generative model, T5-EHR, to the best-performing approaches from the earlier experiments. From the BiLSTM evaluation, Bio-ClinicalBERT combined with BiLSTM emerged as the strongest model, while among standalone transformers, RoBERTa-base-PM-M3-Voc represented the best model.

As shown in Table 5, T5-EHR performed better than both Bio-ClinicalBERT + BiLSTM and RoBERTa, reinforcing the potential of generative models for SBDH classification. To further test whether sequential modeling could enhance T5-EHR, we also evaluated a T5-EHR + BiLSTM configuration. In this case, we observed a similar decline to what was seen with RoBERTa when BiLSTM was added, although the drop in performance was less noticeable. These results suggest that T5-EHR effectively captures sequential dependencies during fine-tuning, making additional sequence modeling unnecessary.

**Table 5:** Comparison of T5-EHR with Prior Best-Performing Models.

| **SBDH** | **Bio-ClinicalBERT + BiLSTM** |  |  | **RoBERTa** |  |  | **T5-EHR** |  |  |
| --- | --- | --- | --- | --- | --- | --- | --- | --- | --- |
|  | *P* | *R* | *F1* | *P* | *R* | *F1* | *P* | *R* | *F1* |
| Alcohol Use | 0.83 | 0.86 | 0.84 | 0.89 | 0.87 | 0.87 | 0.91 | 0.91 | 0.91 |
| Tobacco Use | 0.88 | 0.89 | 0.88 | 0.90 | 0.90 | 0.90 | 0.90 | 0.90 | 0.90 |
| Drug Use | 0.75 | 0.74 | 0.74 | 0.76 | 0.74 | 0.75 | 0.79 | 0.74 | 0.76 |

### Improvement in State-of-the-Art Benchmarks

Finally, we benchmarked our results against the published MIMIC-SBDH study to assess how much improvement our approaches achieved over the state of the art. Table 6 presents a direct comparison of F1 scores across the five SBDH labels: Present, Past, Never, Unsure, and None.

**Table 6:** Comparison of Our Best Approaches Against the MIMIC-SBDH Benchmark (F1 Scores)

| SBDH | [8]’s results |  | Best hybrid | Best Standalone | Our Model |
| --- | --- | --- | --- | --- | --- |
|  | *Bio-ClinicalBERT* | *Best* | *Bio-ClinicalBERT + BiLSTM* | *RoBERTa* | *T5-EHR* |
| Alcohol Use | 0.71 | 0.74* | 0.84 | 0.87 | 0.91 |
| Tobacco Use | 0.75 | 0.75 | 0.88 | 0.90 | 0.90 |
| Drug Use | 0.58 | 0.70* | 0.74 | 0.75 | 0.76 |

\* XGBoost

As shown in Table 6, the results demonstrate clear improvements across all five labels. The combination of class weighting, BiLSTM integration for Bio-ClinicalBERT, and our pretrained T5-EHR contributed to this performance boost. Compared to the benchmark, our models consistently achieved higher F1 scores, highlighting both improved robustness and greater balance across labels. These findings confirm that addressing class imbalance, leveraging model-specific enhancements such as BiLSTM where beneficial, and using generative models can advance the state of the art in SBDH classification. Beyond outperforming prior baselines, our framework demonstrates the value of tailoring model needs and strategies to the unique challenges of clinical NLP tasks.

### Social Determinants of Health Classification

In addition to focusing on behavioral determinants, we also explored the task of social determinants of health classification. We selected T5-EHR and RoBERTa, as both models achieved strong standalone performance without the need for an additional BiLSTM layer. We compared these results directly with those reported in the benchmark study to assess generalizability beyond behavioral determinants. Table 7 summarizes the performance across these additional categories. Notably, the best score for the Environment category in the benchmark was achieved by a Random Forest model rather than a transformer-based method.

**Table 7:** Comparison of SDOHs Results Against the MIMIC-SBDH Benchmark (F1 Scores)

| SBDH | [8]’s results |  |  |  |
| --- | --- | --- | --- | --- |
|  | *Bio-ClinicalBERT* | *Best* | *RoBERTa* | *T5-EHR* |
| Community-Present | 0.96 | 0.96 | 0.99 | 0.99 |
| Community-Absent | 0.91 | 0.91 | 0.94 | 0.96 |
| Education | 0.84 | 0.84 | 0.95 | 0.94 |
| Economics | 0.90 | 0.90 | 0.90 | 0.88 |
| Environment | 0.80 | 0.91* | 0.81 | 0.88 |

\* Random Forest

## Discussion

### Impact of Class Imbalance

The extreme class imbalance affected the models negatively. This imbalance led to a significant bias towards the majority classes, causing the models to perform poorly on the minority classes, as shown in Table 2. In clinical applications, such biases are particularly problematic as they can lead to inaccurate predictions and missed identifications of critical factors that impact patient care. Therefore, we evaluated methods of oversampling, undersampling, and class weights. As shown in Table 2, models benefit from these methods and have better results compared to using the data without performing any such actions.

Implementing these methods helped to mitigate the bias, leading to more balanced model performance across different classes and improving the models’ ability to detect minority class instances.

Also, Figure 4 shows that without using any method, we had zero recall in some classes. This indicates that the models failed to identify any instances of certain classes when class imbalance was not addressed, highlighting the severity of the issue. Figure 4 also shows a difference between no class imbalance methods and class weights and the improvement the model has achieved. The class weights method demonstrated a significant enhancement in recall and overall performance, as it adjusts the loss function to give more importance to the minority classes without altering the dataset. However, it is always ideal not to modify the dataset as much as possible, therefore, the class weights method is the recommended technique to utilize. This method preserves the integrity of the original data while effectively addressing the imbalance, making it a practical and efficient solution for dealing with extreme class imbalance in clinical datasets. By ensuring that the minority classes are adequately represented in the training process, the class weights method enhances the overall reliability and accuracy of the model’s predictions.

**Figure 4:**
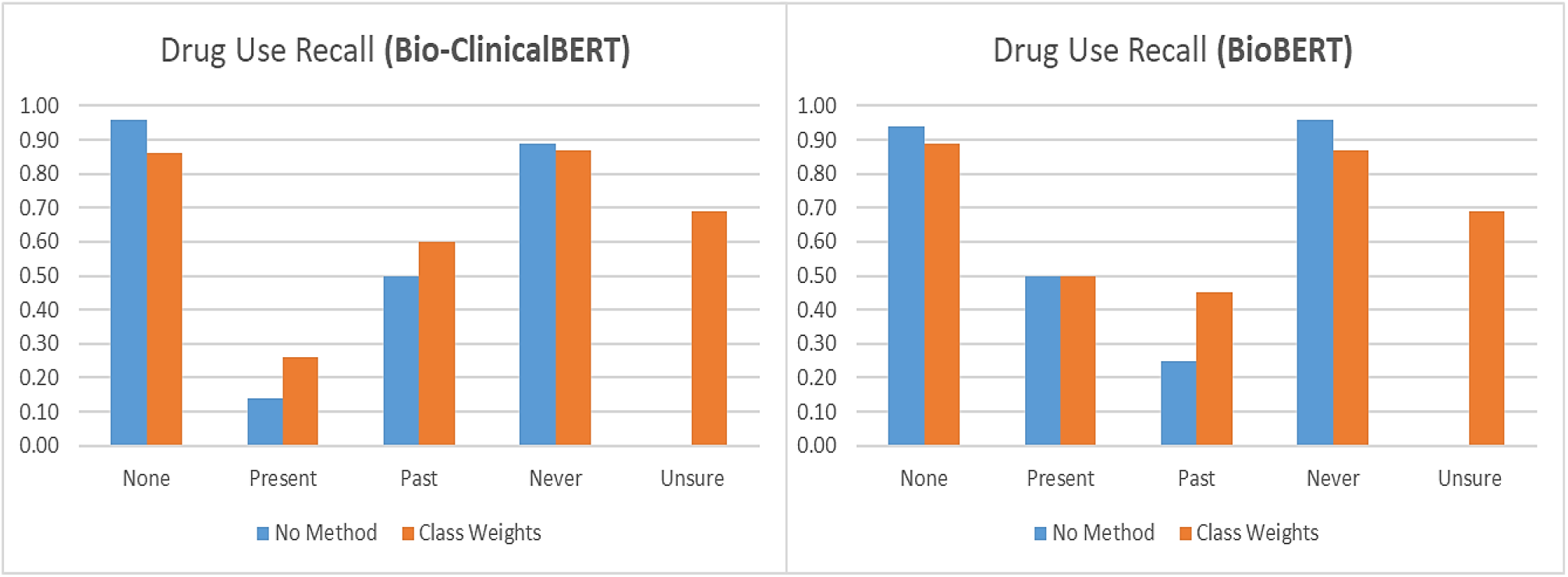
Recall changes when using class weights.

## Results

It is notable that our Bio-Clinical-BERT experiment and [8]’s experiment has slightly different F1 scores results. Such variations are common in machine learning experiments and can be attributed to several factors. First, having a different batch size than [8]’s experiment, which they did not mention in the study, could impact the results. Batch size affects the gradient estimation during training, potentially leading to differences in the model’s learning dynamics and performance. Also, in general, deep learning models, including BERT, often use random initialization for their weights. This random initialization can lead to different convergence paths and, consequently, different final performances during training, introducing variability even when other conditions are controlled. Finally, the training data itself could be different in both experiments. Differences in the specific subset of data used for training can introduce variability in model performance, as the models may be exposed to different examples and patterns. Another key difference is the imbalance-handling strategy. They used oversampling, while we used class weighting.

## Conclusion

Overall, natural language processing (NLP) models represent the most effective technology for extracting and classifying SBDH concepts from clinical narratives. These models can process large volumes of unstructured text data, uncovering critical insights into patients’ social and behavioral determinants of health, which are often not captured in structured data. In this work, we systematically evaluated strategies for addressing class imbalance, investigated the contribution of BiLSTM when combined with transformer embeddings, and explored the role of generative models with T5-EHR. Our results showed that class weighting was the most effective strategy for mitigating imbalance, ensuring fairer performance across all SBDH categories. We also found that BiLSTM integration improved the performance of Bio-ClinicalBERT, BioBERT, and BioMedBERT, while models such as RoBERTa and T5-EHR showed lower performance.

Among all models, our domain-specific generative model, T5-EHR, achieved the strongest overall results, surpassing prior baselines and outperforming the published MIMIC-SBDH benchmark. These findings highlight three key contributions: first, the importance of selecting appropriate strategies for imbalance handling in clinical text classification; second, the conditional value of BiLSTM for enhancing sequential modeling in certain embeddings; and third, the effectiveness of generative models in achieving state-of-the-art performance for SBDH extraction.

## Data Availability

All data produced are available online at the cited work in the paper

https://physionet.org/content/mimiciii/1.4/

https://pmc.ncbi.nlm.nih.gov/articles/PMC8734043/

## Acknowledgment

I would like to express my sincere gratitude to the Saudi Arabian government for their generous funding and support through the PhD scholarship program. This invaluable assistance has enabled me to pursue my research and academic goals. I would also like to extend my heartfelt thanks to my colleagues in the UD NLP lab for their unwavering support and collaboration. Also, this work is partially supported by grant number R35GM141873 from the NIH.

## Footnotes

1 Table 1 provides a description of the labels on tobacco use. The same definition is used in other cases.

## Notes

### Competing Interest Statement

The authors have declared no competing interest.

### Funding Statement

- PhD scholarship program was funded by Saudi Arabian government
- This work is partially supported by grant number R35GM141873 from the NIH.

## References

1. Marmot, M., et al., Closing the gap in a generation: health equity through action on the social determinants of health. Lancet, 2008. 372(9650): p. 1661–9.

2. Chen, E.S., et al., A multi-site content analysis of social history information in clinical notes. AMIA Annu Symp Proc, 2011. 2011: p. 227–36.

3. Alsentzer, E., et al., Publicly available clinical BERT embeddings. arXiv preprint arXiv:1904.03323, 2019.

4. Lee, J., et al., BioBERT: a pre-trained biomedical language representation model for biomedical text mining. Bioinformatics, 2020. 36(4): p. 1234–1240.

5. Gu, Y., et al., Domain-specific language model pretraining for biomedical natural language processing. ACM Transactions on Computing for Healthcare (HEALTH), 2021. 3(1): p. 1–23.

6. Lewis, P., et al. Pretrained language models for biomedical and clinical tasks: understanding and extending the state-of-the-art. in Proceedings of the 3rd clinical natural language processing workshop. 2020.

7. Schuster, M. and K.K. Paliwal, Bidirectional recurrent neural networks. IEEE Transactions on Signal Processing, 1997. 45(11): p. 2673–2681.

8. Ahsan, H., et al., MIMIC-SBDH: A Dataset for Social and Behavioral Determinants of Health. Proc Mach Learn Res, 2021. 149: p. 391–413.

9. Kenton, J.D.M.-W.C. and L.K. Toutanova. Bert: Pre-training of deep bidirectional transformers for language understanding. in Proceedings of naacL-HLT. 2019.

10. Hochreiter, S. and J. Schmidhuber, Long short-term memory. Neural Comput, 1997. 9(8): p. 1735–80.

11. Feller, D.J., et al., Detecting Social and Behavioral Determinants of Health with Structured and Free-Text Clinical Data. Appl Clin Inform, 2020. 11(1): p. 172–181.

12. Stemerman, R., et al., Identification of social determinants of health using multi-label classification of electronic health record clinical notes. JAMIA Open, 2021. 4(3): p. ooaa069.

13. Topaz, M., et al., Extracting Alcohol and Substance Abuse Status from Clinical Notes: The Added Value of Nursing Data. Stud Health Technol Inform, 2019. 264: p. 1056–1060.

14. Yu, Z., et al., A Study of Social and Behavioral Determinants of Health in Lung Cancer Patients Using Transformers-based Natural Language Processing Models. AMIA Annu Symp Proc, 2021. 2021: p. 1225–1233.

15. Liu, Y., et al., Roberta: A robustly optimized bert pretraining approach. arXiv preprint arXiv:1907.11692, 2019.

16. Zhou, L., et al., Using Medical Text Extraction, Reasoning and Mapping System (MTERMS) to process medication information in outpatient clinical notes. AMIA Annu Symp Proc, 2011. 2011: p. 1639–48.

17. Navathe, A.S., et al., Hospital Readmission and Social Risk Factors Identified from Physician Notes. Health Serv Res, 2018. 53(2): p. 1110–1136.

18. Han, S., et al., Classifying social determinants of health from unstructured electronic health records using deep learning-based natural language processing. J Biomed Inform, 2022. 127: p. 103984.

19. Wu, W., et al., Natural language processing to identify social determinants of health in Alzheimer’s disease and related dementia from electronic health records. Health Serv Res, 2023. 58(6): p. 1292–1302.

20. Richie, R., et al., Extracting social determinants of health events with transformer-based multitask, multilabel named entity recognition. J Am Med Inform Assoc, 2023. 30(8): p. 1379–1388.

21. Lybarger, K., et al., Leveraging natural language processing to augment structured social determinants of health data in the electronic health record. J Am Med Inform Assoc, 2023. 30(8): p. 1389–1397.

22. Gou, Z. and Y. Li, Integrating BERT Embeddings and BiLSTM for Emotion Analysis of Dialogue. Computational Intelligence and Neuroscience, 2023. 2023(1): p. 6618452.

23. Jiang, X., et al., Research on sentiment classification for netizens based on the BERT-BiLSTM-TextCNN model. PeerJ Computer Science, 2022. 8: p. e1005.

24. Bello, A., S.-C. Ng, and M.-F. Leung, A BERT framework to sentiment analysis of tweets. Sensors, 2023. 23(1): p. 506.

25. Johnson, A.E., et al., MIMIC-III, a freely accessible critical care database. Sci Data, 2016. 3: p. 160035.

26. Hettiarachchi, H., et al. DAAI at CASE 2021 task 1: Transformer-based multilingual socio-political and crisis event detection. in Proceedings of the 4th Workshop on Challenges and Applications of Automated Extraction of Socio-political Events from Text (CASE 2021). 2021.

